# Insights into Professional Preferences and Rationale for Surgical Sequencing in Managing Hip-Spine Syndrome

**DOI:** 10.1101/2024.04.13.24305764

**Authors:** Carolina Breuning, Xinggui Tian, Jens Goronzy, Klaus-Peter Günther, Uwe Platz, Franziska Beyer, Alexander Carl Disch, Paul F. Lachiewicz, Ning Liu, Stuart B. Goodman, Kirkham B. Wood, Stefan Zwingenberger

## Abstract

**Background:** The optimal sequence of hip and spine surgeries for hip-spine syndrome management remains a contentious issue. This study aimed to investigate the preferences and rationale behind the sequence of surgical treatments across various hip-spine syndrome scenarios among potential specialist interviewees.

**Methods:** A questionnaire survey, featuring five fictional clinical presentations encompassing symptomatic hip osteoarthritis and diverse spinal pathologies, was conducted among German hip and spine surgeons, alongside conducting a cross-national comparison with previous US survey.

**Results:** German hip and spine surgeons preferred spine-first surgery in hip-spine syndrome scenarios involving neurological deficits, while preferred hip-first in scenarios without such deficits. In contrast, US surgeons demonstrated differing sequencing patterns, highlighting differences between nations. Notably, distinct surgical order preferences were observed among different specialists. Surgeons’ decision-making was primarily influenced by symptom severity and urgency, spine-pelvis-hip biomechanics, and treatment efficacy.

**Conclusion:** The sequence of hip and spine surgery in various hip-spine syndrome scenarios has different preference patterns, with professional preferences and cross-nation differences, and is guided by the multifaceted considerations involved in surgical decision-making.

## 1. Introduction

The hip-spine syndrome was first described by Offierski et al. in 1983, who defined this condition as the presence of concomitant hip and spine pathology and categorized it as “simple”, “complex”, “secondary” and “misdiagnosed” (1). In simple categories, the prioritization of surgical treatment may be clear due to straight forward of symptoms; however, in complex cases, the prioritization of surgical treatment for the hip-spine syndrome is challenging due to the substantial overlap of symptomatology. Moreover, there is currently no consensus on which pathology should be addressed first (1, 2). Previous studies mainly focused on the effects of the sequence of total hip arthroplasty (THA) for hip disease and spinal fusion surgery for spinal disease on each other’s outcomes (3–7), while few studies have focused on the patient-centered benefits based on the different clinical presentations for the hip-spine syndrome. Different scenarios of the hip-spine syndrome may benefit differently from the surgical sequences, such as ankylosing spondylitis, spinal stenosis, and degenerative spondylolisthesis with concomitant hip OA (8, 9). Therefore, it is clinically important to investigate the patient-centered treatment sequence of different diseases.

In the clinical setting, joint surgeons and spine surgeons must face whether patients with symptoms in the hip and lumbar spine who require surgical treatment should have the hip or the spine operation first. Currently, the surgical sequence of treatment for hip-spine syndrome is controversial from the perspective of spine or arthroplasty surgeons (10, 11). There are currently no explicit guidelines for the hip-spine syndrome, which is why an interprofessional consultation with hip surgeons (HS) and spine surgeons (SS) is clinically important. A collaborative research team from Stanford University consulted HS and SS across the USA on the preferred treatment sequence of five fictional scenarios in patients with hip OA and common degenerative lumbar diseases and found that there are still controversies in some clinical situations, even among experienced surgeons (12). Due to the different training patterns of orthopedic surgeons in the USA and Germany (13), this may lead to different preferences for the surgical sequence in patients with the hip-spine syndrome between the two countries and warrants further investigation. Spine surgery is also performed in neurosurgery, in addition to orthopedic surgery in the USA and Germany, while differences in structured surgical residency training during neurosurgery and orthopedic residency in Germany (14) the USA (15) and the aforementioned differences between two countries (13, 16, 17) may also result in different treatment options of surgical sequence.

Therefore, the aim of this study was to investigate the interprofessional choice of surgical sequence for different hip-spine syndrome scenarios between German HS and SS (orthopaedic spine surgeon (OSS) and neurosurgical spine surgeons (NSS)), and to compare the findings with the same questionnaire previously conducted in the USA to raise awareness of the complex treatment algorithm for patients and use it for “patient-centered shared decision-making”. The hypothesis of this study was, that surgeons may have different surgical order preferences for different hip-spine syndrome scenarios, based on professional preferences between different specialists, and cross-nation differences between German and US surgeons.

## 2. Methods

### 2.1 Questionnaire design

An online questionnaire in English with five fictional hip-spine syndrome scenarios was previously designed by the Stanford University research team, including clinical history, current symptoms, further diagnosis and related images of the hip and spine (12). Five fictional hip-spine syndrome scenarios describe symptomatic hip OA with five common degenerative spinal diseases: 1) degenerative lumbar spinal stenosis with neurogenic claudication, 2) degenerative lumbar spondylolisthesis with leg pain, 3) a single-level lumbar disc herniation with weakness in the leg, 4) lumbar scoliosis with the sagittal imbalance and back pain and 5) thoracolumbar disc herniation with signs of myelopathy. The surgeons’ preferences for the order of treatment were collected for each scenario, and to provide a reason for their choice in free-text comments. Additionally, they had to select which kind of hip articulation (standard size head, large head >32mm, dual mobility implant or constrained liner) they would choose if they chose THA first **(Figure S1)**. The English questionnaire was translated into German (by two senior German physicians and was subsequently proofread by another senior physician) in this study and sent to German surgeons using an online survey platform “LimeSurvey” (LimeSurvey GmbH, Hamburg, Germany). The reporting of this study conforms to the Checklist for Reporting Results of Internet E-Surveys (CHERRIES) statement (18) **(Table. S1)**.

### 2.2 Survey

This survey was then sent to 2500 members of the “German Spine Society” (OSS and NSS) on March 26^th^, 2021, and 883 members of the “German Society for Joint Replacement” (Arbeitsgemeinschaft für Endoprothetik, mostly HS) on April 8^th^, 2021 *via* mail by the representatives of the associations. The choices of different German specialists in each scenario, and the relation between the year of experience and choice were analyzed. Text-mining was used to analyze the free-text comments and to identify the most frequently used words. It was conducted with R version 4.0.4 using word frequency packages tidyverse (version 1.3.1), tidytext (version 0.3.2), wildyr, igraph and ggraph. Then, the list of the words was sorted alphabetically in Excel to summarize words with an equal meaning in main groups. For the transfer to Excel the packages readxl (version 1.3.1) and xlsx (version 0.6.5) were used. The density for each word was calculated as the number of the specific word divided by the total word count. The comments were summarized into key points explaining the most commonly used words. The results of the survey of surgeons in Germany were further compared, point-to-point, with the same survey of surgeons in the USA previously done by Stanford University (12). In particular, the surgeons participating in this survey in Germany were HS from the German Society for Joint Replacement and SS from German Spine Society, including NSS and OSS, while the surgeons participating in the survey in the USA were HS from the North American Hip Society and SS from the Scoliosis Research Society, not further divided into NSS and OSS (12). Therefore, merging the German NSS and OSS into an overall SS was performed, when comparing the results of the surveys of SS between German and USA.

### 2.3 Statistical analysis

Data were presented as percentages or mean (range) in this study using SPSS version 20 (SPSS, Inc., Chicago, IL, USA) and choices between two groups were compared using the chi-square test. P ≤ 0.05 was considered statistically significant.

## 3. Results

### 3.1 Participants

7.02% (62/883) of the German HS and 3.88% (97/2500) of the SS responded to the survey. In total, 159 German surgeons participated in this study, consisting of 38.3% HS, 24.1% NSS, and 35.8% OSS. The USA survey received 88 surgeon responses to the survey, 46% (51/110) of HS responded and 37% (37/101) of SS responded (12). The US survey had a relatively high response rate compared to the German survey, which may be due to the different sample selection methods. The US adopted a sample survey method while Germany selected a full-sample survey.

### 3.2 Preference patterns of surgical sequence

**Table 1** presents the outcomes of the survey detailing the preferred treatment sequences selected by German and American surgeons in response to five fictional clinical scenarios. Overall, a consistent pattern of surgical preference was achieved among German surgeons, in which spine-first preferred in scenarios 1, 3, and 5 and hip-first preferred in scenarios 2 and 4. Although SS and HS have different percentage preference for surgical sequence, none of the five conditions were statistically different. In scenario 1, SS had a higher proportion of spine-first than HS, which was not statistically significant, while NSS significantly preferred spine-first to OSS. In scenarios 2 and 4, the difference of selection ratio between SS and HS was not significant, but OSS preferred hip over NSS at a significantly higher rate in scenario 2. In scenario 3, either the OSS or the NSS favored spine-first over the HS. In scenario 5, spine-first was preferred by most of HS and SS (OSS and NSS) in Germany.

**Table 1.** The preferred sequence of treatment chosen by German and American surgeons in five fictional clinical scenarios. U.S. data was from one published article by Stanford University (*12*).

| Germany |  |  |  |  | USA<sup>1</sup> |  |  |  |  |
| --- | --- | --- | --- | --- | --- | --- | --- | --- | --- |
| Scenario | Hip-first (%) | Spine-first (%) | No preference (%) | χ-value; p-value | Scenario | Hip-first (%) | Spine-first (%) | No preference (%) | χ-value; p-value |
| **Case 1<sup>a</sup>** |  |  |  | 2.850; 0.240 | **Case 1<sup>a</sup>** |  |  |  | 0.532; 0.466 |
| Hip surgeon | 35.5 | 53.2 | 11.3 | (Hip vs. Spine surgeon) | Hip surgeon | 59 | 33 | 8 | (Hip vs. Spine surgeon) |
| Spine surgeon | 30.9 | 63.9 | 5.2 |  | Spine surgeon | 49 | 46 | 5 |  |
| NSS | 7.7 | 87.2 | 5.1 | 16.908; <0.001*<br>(NSS vs. OSS) |  |  |  |  |  |
| OSS | 46.6 | 48.3 | 5.2 |  |  |  |  |  |  |
| **Case 2<sup>b</sup>** |  |  |  | 4.584; 0.101 | **Case 2<sup>b</sup>** |  |  |  | 0.001; 1.000 |
| Hip surgeon | 88.7 | 8.1 | 3.2 | (Hip vs. Spine surgeon) | Hip surgeon | 73 | 18 | 10 | (Hip vs. Spine surgeon) |
| Spine surgeon | 79.4 | 7.2 | 13.4 |  | Spine surgeon | 70 | 8 | 22 |  |
| NSS | 61.5 | 12.8 | 25.6 | 12.744; 0.002*<br>(NSS vs. OSS) |  |  |  |  |  |
| OSS | 91.4 | 3.4 | 5.2 |  |  |  |  |  |  |
| **Case 3<sup>c</sup>** |  |  |  | 2.027; 0.363 | **Case 3<sup>c</sup>** |  |  |  | 6.259; 0.012* |
| Hip surgeon | 41.9 | 48.4 | 9.7 | (Hip vs. Spine surgeon) | Hip surgeon | 47 | 45 | 8 | (Hip vs. Spine surgeon) |
| Spine surgeon | 32 | 59.8 | 8.2 |  | Spine surgeon | 19 | 73 | 8 |  |
| NSS | 33.3 | 56.4 | 10.3 | 0.483; 0.786<br>(NSS vs. OSS) |  |  |  |  |  |
| OSS | 31 | 62.1 | 6.9 |  |  |  |  |  |  |
| **Case 4<sup>d</sup>** |  |  |  | 0.095; 0.954 | **Case 4<sup>d</sup>** |  |  |  | 7.552; 0.006* |
| Hip surgeon | 71 | 17.7 | 11.3 | (Hip vs. Spine surgeon) | Hip surgeon | 47 | 47 | 6 | (Hip vs. Spine surgeon) |
| Spine surgeon | 73.2 | 16.5 | 10.3 |  | Spine surgeon | 78 | 11 | 11 |  |
| NSS | 71.8 | 15.4 | 12.8 | 0.465; 0.792<br>(NSS vs. OSS) |  |  |  |  |  |
| OSS | 74.1 | 17.2 | 8.6 |  |  |  |  |  |  |
| **Case 5<sup>e</sup>** |  |  |  | 4.049; 0.132 | **Case 5<sup>e</sup>** |  |  |  | 2.234; 0.071 |
| Hip surgeon | 6.5 | 88.7 | 4.8 | (Hip vs. Spine surgeon) | Hip surgeon | 10 | 86 | 4 | (Hip vs. Spine surgeon) |
| Spine surgeon | 1 | 95.9 | 3.1 |  | Spine surgeon | 0 | 97 | 3 |  |
| NSS | 0 | 97.4 | 2.6 | 0.748; 0.688<br>(NSS vs. OSS) |  |  |  |  |  |
| OSS | 1.7 | 94.8 | 3.4 |  |  |  |  |  |  |

\*Statistically significant
<sup>a</sup> Lumbar canal stenosis with neurogenic claudication combined with osteoarthritis of the hip
<sup>b</sup> Degenerative lumbar spondylolisthesis with radicular leg pain combined with osteoarthritis of the hip
<sup>c</sup> Lumbar disc herniation with muscle strength weakness combined with osteoarthritis of the hip
<sup>d</sup> Scoliosis with back pain combined with osteoarthritis of the hip
<sup>e</sup> Thoracolumbar disc herniation with myelopathy combined with osteoarthritis of the hip
“NSS” and “OSS” indicated the neurosurgical spine surgeon and orthopaedic spine surgeon in the survey of
Germany, respectively.

A further German-American comparison found that the German-US preference patterns were consistent in scenario 2 (HS and SS), scenario 5 (HS and SS), scenario 3 (SS), and 4 (SS), but reversed in scenario 1 (HS and SS). The preferences of HS in the USA were debated in scenarios 3 and 4. In scenario 1, more American surgeons (HS and SS) preferred to hip-first, whereas more German surgeons (HS and SS) preferred to spine-first. In scenario 2, both American and German surgeons prioritized the hip-first. In scenario 3, German surgeons as well as SS in the USA preferred spine-first, while HS in the USA showed a debate about the preference of the surgical sequences. In scenario 4, German surgeons and American SS prefer hip-first, while American HS had no clear preference for order of surgery. In scenario 5, both American and Germany surgeons (HS and SS) preferred to spine-first.

### 3.3 Rationale for order of treatment choices

The most frequently used words from the free-text comments are shown in **Figure 1**. In scenario 1, German surgeons preferred hip-first due to considering “*hip*” to be more “symptomatic” and therefore increasing lordosis of the spine and recommending THA first, and while spine-first was preferred because spinal “*stenosis*” was considered more “symptomatic”. American surgeons preferred hip-first considered that THA can improve spine-pelvic biomechanics and thereby spinal symptoms “*relief*”, while those who chose spine-first believed that untreated neurogenic claudication may hinder the “*recovery*” of THA. In scenario 2, German surgeons preferred THA first because of the severe “*symptomatic coxarthrosis*” in the “*right hip*”. After THA, the patient’s mobility and spinal alignment would improve significantly, resulting in improved spinal symptoms, and also conducive to better rehabilitation training for subsequent spinal surgery. The American surgeons considered hip problems to be “*serious*” or “*more severe*” than radicular leg pain in the absence of neuromotor deficits, and THA was also thought to provide more predictable pain “*relief*” than “*spinal fusion*”, leading to hip-first preferred. In scenario 3, some German surgeons prioritized hip-first, as “*femoral head necrosis*” is an urgent “*indication*” for “hip”, while those who choose spine-first considered the neurological symptoms from the “*disc*” herniation, such as muscle strength weakness and paralysis, to be signs of impending nerve damage, therefore spine-first preferred. Similarly, most American surgeons prioritized spine-first because muscle “*weakness*” is a “*neurological deficit*”, and “discectomy” is relatively simple for an extruded “*disc*”. In scenario 4, most German surgeons considered that “*scoliosis*” represents a chronic event compared to severe hip symptoms, whereas THA will provide significant pain relief and has a positive effect on chronic back pain, while some surgeons preferred spine-first to correct “*sagittal*” imbalance of spine. Many US THA surgeons commented that balancing the spine first was important to optimize the “*position*” of subsequent THA components as this allows for “changes” in the spine-pelvic alignment due to spinal surgery, while many THA surgeons still treat the hip-first, assuming the patient’s “*nerves*” were intact, and spine surgery was not urgent. Prioritizing hip-first was a practical consideration for most US spine surgeons, as THA is an “*easier*” procedure with more “*predictable*” results and faster than scoliosis surgery recovery. In scenario 5, most of German surgeons would treat spine-first showing signs of “*myelopathy*” and spinal cord “*compression*” to prevent irreversible neurological deficits. Likewise, “*myelopathy*” caused by “*compression of the spinal cord*” by the American surgeons was considered more urgent than the hip OA (12).

**Figure 1.**
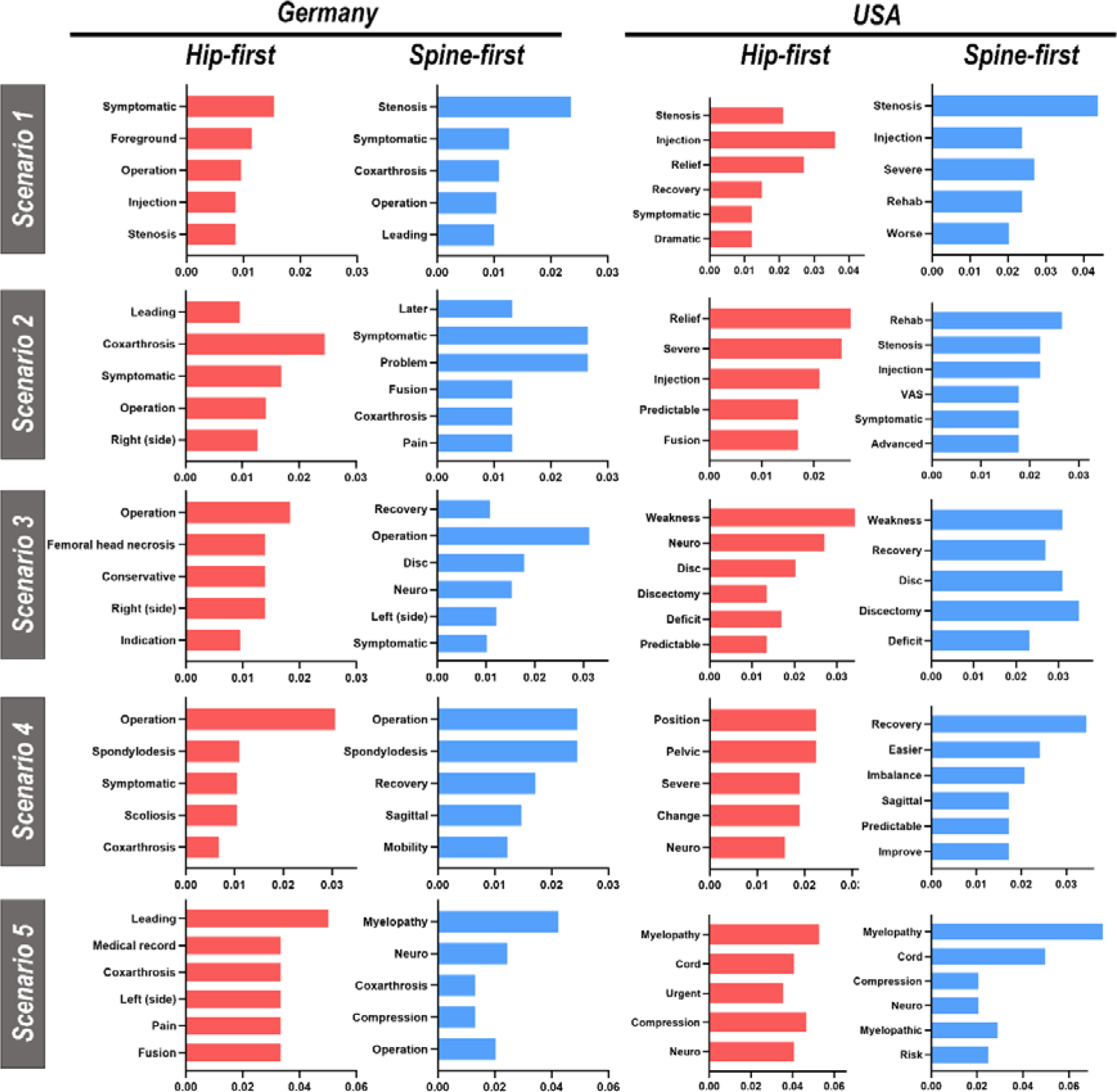
The most frequently used words in the surgeons’ comments for the five scenarios. U.S. data was from one published article by Stanford University (12).

### 3.4 Impact of years of experience on treatment choices

The majority of German HS and NSS had between 11 and 20 years of experience (35.5% and 43.6%, respectively), while most OSS had between 0 to 10 years of experience (39.7%) **(Figure 2)**. On average, German surgeons had 18.3 (2–36) years of experience, with HS, NSS, and OSS having an average of 20.5 (3–36), 20.1 (2–34), and 14.8 (2–34) years of experience, respectively. Average years of experience post-training for HS was 30.8 (14-60 years) and 23.4 (5-34 years) for SS in the American study (12).

**Figure 2.**
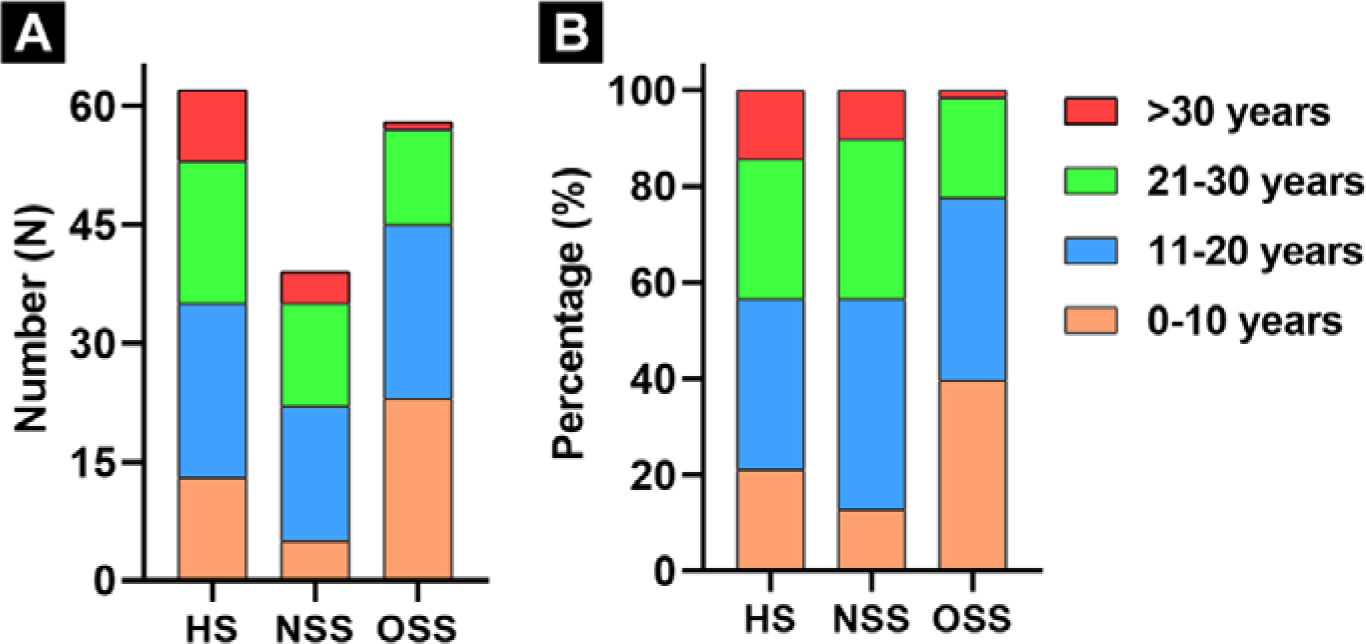
The number **(A)** and percentage **(B)** of participants in three different German specialties at different experience years after surgical training. “HS”, “NSS” and “OSS” indicated the hip surgeon and neurosurgical spine surgeon and orthopaedic spine surgeon, respectively.

Analysis of the association between years of experience of German surgeons and treatment priorities showed the years of experience of the participants did not significantly affect the general trend in treatment choice preference, as manifested by a preference for spine-first in scenario 1, 3, and 5 and hip-first in scenario 2 and 4 in any year of experience levels, although the proportions varied at different experience levels **(Table 2)**. Some patterns were noticed in the scenario 1 and 5 as the surgeons became more experienced in Germany. The “Spine-first” proportion increased with years of experience in scenario 1, while the proportion of “Hip-first” gradually increased from 0 to about 7%, as the surgeon became more experienced in scenario 5, which indicated that the degree of attention to neurological deficit symptoms may vary with surgeon experience. The study cannot safely draw conclusions about the impact of years of experience on different specialists due to the small sample size at different experience levels **(Table. S2-6)**.

**Table 2.** The surgeons’ preferences for treatment order at different experience levels in five fictional clinical scenarios in Germany.

| Years of experience |  |  |  |  |
| --- | --- | --- | --- | --- |
| Scenario | Hip-first (%) | Spine-first (%) | No preference (%) | χ-value; p-value |
| **Case 1<sup>a</sup>** |  |  |  | 6.240; 0.397 |
| 0-10 years | 43.9 | 51.2 | 4.9 |  |
| 11-20 years | 29.5 | 60.7 | 9.8 |  |
| 21-30 years | 32.6 | 62.8 | 4.7 |  |
| > 30 years | 14.3 | 71.4 | 14.3 |  |
| **Case 2<sup>b</sup>** |  |  |  | 6.462; 0.373 |
| 0-10 years | 87.8 | 7.3 | 4.9 |  |
| 11-20 years | 86.9 | 6.6 | 6.6 |  |
| 21-30 years | 72.1 | 9.3 | 18.6 |  |
| > 30 years | 85.7 | 7.1 | 7.1 |  |
| **Case 3<sup>c</sup>** |  |  |  | 1.867; 0.932 |
| 0-10 years | 34.1 | 56.1 | 9.8 |  |
| 11-20 years | 39.3 | 50.8 | 9.8 |  |
| 21-30 years | 30.2 | 62.8 | 7 |  |
| > 30 years | 42.9 | 50 | 7.1 |  |
| **Case 4<sup>d</sup>** |  |  |  | 4.889; 0.558 |
| 0-10 years | 68.3 | 14.6 | 17.1 |  |
| 11-20 years | 75.4 | 16.4 | 8.2 |  |
| 21-30 years | 72.1 | 16.3 | 11.6 |  |
| > 30 years | 71.4 | 28.6 | 0 |  |
| **Case 5<sup>e</sup>** |  |  |  | 5.184; 0.520 |
| 0-10 years | 0 | 92.7 | 7.3 |  |
| 11-20 years | 3.3 | 95.1 | 1.6 |  |
| 21-30 years | 4.7 | 93 | 2.3 |  |
| > 30 years | 7.1 | 85.7 | 7.1 |  |

\*Statistically significant
<sup>a</sup> Lumbar canal stenosis with neurogenic claudication combined with osteoarthritis of the hip
<sup>b</sup> Degenerative lumbar spondylolisthesis with radicular leg pain combined with osteoarthritis of the hip
<sup>c</sup> Lumbar disc herniation with muscle strength weakness combined with osteoarthritis of the hip
<sup>d</sup> Scoliosis with back pain combined with osteoarthritis of the hip
<sup>e</sup> Thoracolumbar disc herniation with myelopathy combined with osteoarthritis of the hip

### 3.5 Choice of implant in THA

Surgeons had to specify the articulation type in THA if they preferred hip-first and NSS were excluded due to professional restrictions. German surgeons mostly chose components with standard-size head in scenario 2, 3, 4, and 5, except for scenario 1 where the selection of standard-size head was comparable to that of large head ratio. In contrast, the majority of US surgeons preferred the large head in all scenarios and chose dual mobility more often than German surgeons. In scenario 4, the proportion of choosing dual mobility and large head was significantly higher than other groups to reduce dislocation risk in both surveys(12). (**Figure 3)**

**Figure 3.**
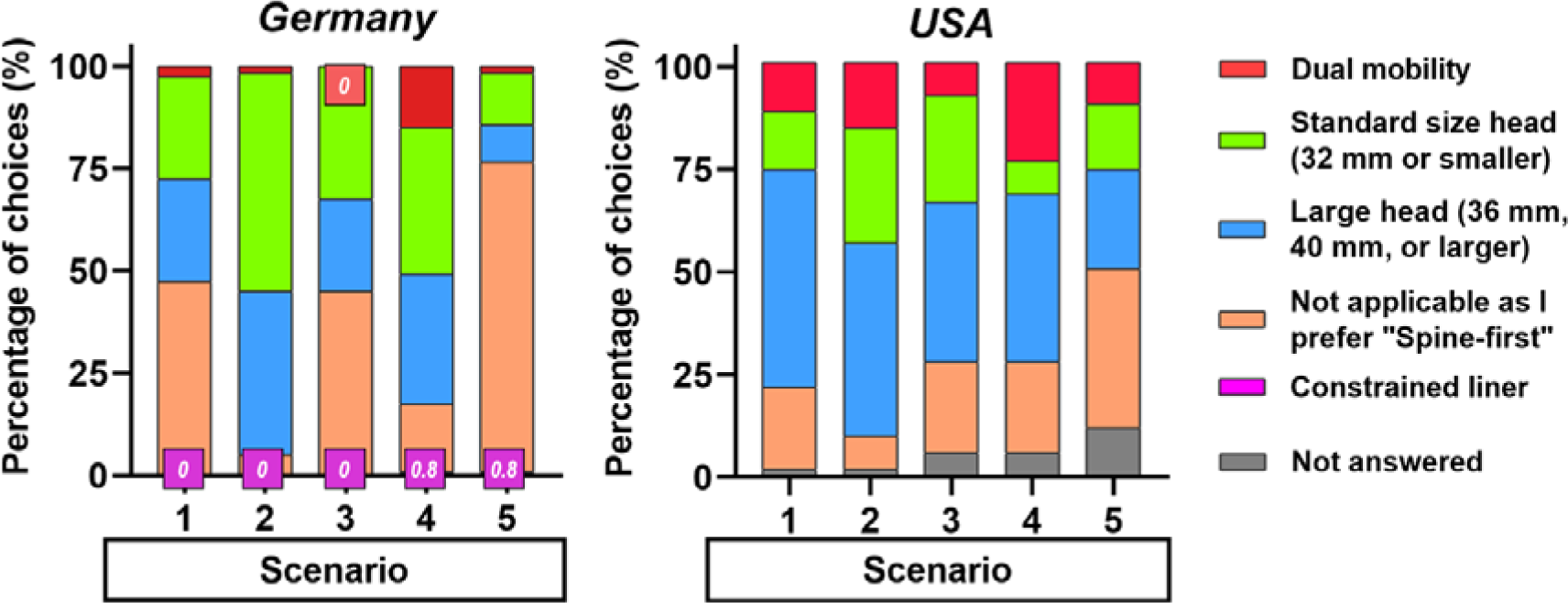
German and American surgeons’ choice of hip articulation type for different scenarios. U.S. data was from one published article by Stanford University (12). All participating German hip surgeons and orthopedic spine surgeons answered this question and none of the participating US surgeons chose a constrained liner option.

## 4. Discussion

The focus of this study was to investigate disease-based treatment sequence preferences in different hip-spine syndrome scenarios at potential visiting specialists through a questionnaire of establishing fictional scenarios and a possible cross-nation comparison between Germany and previous American study. The hypothesis of this study was verified that the sequence of hip and spine surgery in different hip-spine syndrome scenarios has different preference patterns, with professional preferences and cross-nation differences. Moreover, the survey also found that the choice of surgical sequence preference was mainly affected by the severity of neurological deficit symptoms, spine-pelvic-hip biomechanics, and surgery-related factors.

Hip-spine syndrome was first described and classified in 1983 by Offierski et al, but the classification system cannot provide guidance for the order of hip and spine surgery (1). Due to overlapping symptoms, such patients tend to be referred between hip and spine specialists (2). In this study, the investigation of patient-centered shared decision-making among potential visiting specialists showed that German surgeons prioritize spine-first for neurogenic deficiency spine disease with hip OA, otherwise prefer hip-first. Similarly, surgical sequence preference among US surgeons also showed a pattern associated with neurological deficits, particularly for SS, as indicated by a preference for spine-first in scenario 5 with signs of myelopathy and a preference for hip-first in scenario 2 without neurological deficits, and a preference of SS for spine-first in scenario 3 of chronic neurological impairment. The difference was that the American surgeons seem to be less concerned about chronic neurological deficits than German surgeons, especially the American HS, who showed a preference for hip-first in scenario 1, and there were also controversies within the American HS in scenarios 3 and 4, while SS preferred to treat lumbar scoliosis first before hip OA. This demonstrated that the sequence of hip and spine surgery in different hip-spine syndrome scenarios has different preference patterns and cross-nation differences, which suggested that the selection of surgical sequence in different hip-spine syndrome scenarios may require individualized treatment. The percentage of American and Germany SS, especially Germany NSS, who preferred spine-first was higher than HS in cases of neurological deficit, and the percentage of HS who preferred hip-first was higher than SS in all scenarios except accompanying scoliosis, and preferred spine-first was higher than SS in cases with lumbar scoliosis in scenario 4, which indicated the sequence of surgical choice has professional preferences. It is not clear whether the professional preference is due to the lack of understanding of the hip-spine syndrome resulting from the differences in education or training, so this study cannot guide current medical training, but this study potentially points to the importance of comprehensive knowledge and multidisciplinary consultation of both joint and spine surgeons for patients with complex joint and spine diseases. The present study showed that surgeons’ choice of surgical sequence was influenced by many factors. First, the severity of neurological deficits is the most important factor affecting decision-making. Spine-first was preferred by all German and American surgeons when signs of myelopathy derived from thoracolumbar disc herniation are present (scenario 5), which is consistent with previous views among HS and SS (10, 11, 19). Patients showing signs of myelopathy may benefit from early surgery, whereas prolonged preoperative symptom duration may lead to poor outcomes (20). Neurogenic claudication is the result of nerve root ischemia or mechanical compression (21). German surgeons were more concerned with neurogenic claudication than US surgeons and tended to perform spine-first, while US surgeons tended to perform hip-first, although American SS were more concerned with neurogenic claudication than HS (scenario 1). Weakness in the leg is thought to be related to mechanical compression and inflammation of nearby nerve roots and dorsal root ganglia caused by lumbar disc herniation (22). It was observed that German and American SS and German HS were more concerned about the nerve root deficit caused by lumbar disc herniation, while American HS were less concerned (scenario 3). In the case of degenerative spondylolisthesis with leg pain, all surgeons preferred to perform hip-first (scenario 2), as participants considered hip symptoms to be more severe than radicular leg pain in the absence of neurological deficits. In contrast to severe hip symptoms, scoliosis is a chronic event without neurological impairment (scenario 4), so most German surgeons, American SS and half of the American HS believe that spine disease is not an emergency and prioritizing treatment of the hip is a practical consideration.

Surgeons’ decisions are also influenced by the hip-spine-pelvic biomechanical interaction. Hip-spine syndrome involves the interplay of the hip and spine pathology, symptoms, and biomechanics (19). The features of degenerative spine disease and advanced hip disease mean increased stiffness in both the spine and hip joints (19). Spinal fusion surgery makes the spine stiffer, while THA surgery makes the hip more flexible (19). Therefore, most American and German surgeons in scenario 2, and many German and American HS in scenario 1 consider that THA will improve spine-pelvic biomechanics which can relieve spinal disease symptoms and therefore choose THA first. Also, some German SS believed that spinal fusion could improve femoral head position and gait pattern in scenario 1, thus spine-first is recommended. Nearly half of the American HS believed that balancing the spine first of lumbar scoliosis with the sagittal imbalance in scenario 4 could optimize the position of subsequent THA components. One review on hip-spine syndrome also recommended that in patients with severe spinal deformity complicated by hip abnormalities, correcting the spinal deformity first has the potential to reduce hip dislocation after THA due to correct acetabular positioning (2). Considering the biomechanical interaction of the spine-pelvic-hip, previous studies recommend higher stability implants in those at high risk of hip dislocation (23), including the use of larger femoral heads or dual-mobility constructs (3, 4, 24). American and German surgeons chose dual-mobility components in scenario 4 at a higher rate than in other cases, which may be due to lumbar scoliosis with the sagittal imbalance leading to sagittal compensation of the pelvis and reduced loading area of the acetabulum, increasing the risk of dislocation (25). Moreover, the optimal position of the cup to accommodate pelvic parameters and limit impact may lie outside the classical parameters of the safety zone in the case of spinal deformity (26). American HS prefer larger femoral heads and dual-mobility cups (12), while German surgeons prefer the standard-size femoral heads. The reason for this discrepancy between American and German surgeons is unknown and requires further investigation.

In addition, surgery-related factors like difficulty and therapeutic effect will affect the decisions of the order of specific operations made by surgeons. Currently, advances in minimally invasive techniques for THA and spinal minimally invasive procedures have allowed patients to recover quicker and more efficaciously with fewer complications (27), and spine surgery is associated with a higher complication rate compared with hip surgery, and also varies among different spine surgical techniques (10), which may also affect the choice of surgical sequence. Discectomy is simpler than THA, thus favoring spine-first in scenario 3, while THA is considered to be easier than corrective surgery for scoliosis with more predictable results and faster recovery, so hip-first is preferred in scenario 4 by some US surgeons. Minimally invasive surgical techniques often have a steep learning curve. However, whether the difference in the treatment sequence caused by the years of experience is related to the improvement of technology or a deeper understanding of diseases cannot be fully explained from the data obtained in this study.

## 5. Limitations

This study is a survey of fictional clinical cases that puts the patients at the center of decision-making, providing an overview and differences of current views on this topic in the USA and German; however patient preferences and socioeconomic status were not included. Although this study presents five common hip-spine syndrome cases, the clinical situation is often more complex and may not be applicable in other situations; thus, an individualized treatment plan is preferred for a specific patient, and the treatment of patients with the hip-spine syndrome should be based on a full understanding of the disease. The purpose of this study is to inform readers as appropriate in practice and future research, and not to “guide” current practice. Regrettably, the response rate of this survey was low, which may lead to the bias in the results. Especially, the heterogeneous distribution of years of experience post-surgical training may influence the findings, thus no conclusions can be drawn relating the sequence of surgical treatment and years of experience. The relatively large total sample size in both serveries can still provide reference information, and further prospective observation or intervention studies might help further confirm our results and provide more detailed information.

## 6. Conclusion

German surgeons prioritize spine-first for spinal disorders with neurological deficits of hip-spine syndrome, otherwise opting for hip-first. German-US preference patterns were consistent in scenario 2 (HS and SS), scenario 5 (HS and SS), scenario 3 (SS), and 4 (SS), but reversed in scenario 1 (HS and SS). The preferences of HS in the USA were debated in scenarios 3 and 4. The percentage of American and Germany SS, especially Germany NSS, who preferred spine-first, was higher than HS in case of neurological deficit, and the percentage of HS who preferred hip-first was higher than SS in all scenarios except accompanying scoliosis, and preferred spine-first was higher than SS with lumbar scoliosis in scenario 4. The surgeons’ preference was primarily influenced by the severity and time urgency of symptoms, spine-pelvis-hip biomechanics, and ease and therapeutic effect of hip and spine surgery. In summary, the sequence of hip and spine surgery in different hip-spine syndrome scenarios has different preference patterns, with professional preferences and cross-nation differences, affected by many factors including disease and treatment regimen.

## Take home message

1. The sequence of surgery for hip-spine syndrome should prioritize patient-centered approaches, with preferences varying based on individual scenarios.
2. Surgical sequencing decisions for hip-spine syndrome exhibit professional preferences and cross-nation differences.
3. Surgeons’ decisions are influenced by factors such as symptom severity and urgency, as well as considerations related to spine-pelvis-hip biomechanics and treatment efficacy.

## Declarations

### Ethics approval and consent to participate

Ethical approval was waived by the local Ethics Committee of Technical University of Dresden in view of the nature of the research study (No. EK45012019).

### Consent for participants

To protect participant rights, we ensured the provision of informed consent during the distribution of the questionnaire via email. Participants, including spine surgeons and hip surgeons, were provided with comprehensive information regarding the study’s subject, the challenges faced by surgeons, and the study’s objectives and implementation. It was explicitly communicated that the study’s origin was a questionnaire created by Professors Kirkham Wood and Stuart B. Goodman of Stanford University. The survey, which took approximately 5-10 minutes, was offered to participants free of charge, with no anticipated risks or immediate benefits. After completion, the research findings will undergo a rigorous scientific evaluation, presentation, and eventual publication. Regarding participant rights, individuals, upon reviewing the informed consent and deciding to participate in the study, affirmed the voluntary nature of their involvement. They retained the right to withdraw their consent in written form at any time. All collected data will be subject to anonymous evaluation. Additionally, the informed consent form included our contact information and details of the person in charge.

### Copyright statement

The version published in this journal is the only one authorized by the authors.

### Author Contributions

Conceptualization, C.B., P.F.L., N.L., S.B.G., K.B.W., S.Z.; methodology, C.B., X.T., F.B., N.L., S.B.G, S.Z.; software, C.B., X.T., F.B., N.L.; validation, C.B., X.T., S.Z.; formal analysis, C.B., X.T., F.B.; investigation, C.B., S.Z.; resources, J.G., K.P.G., A.C.D, S.B.G, K.B.W.; data curation, C.B., X.T., S.Z.; writing—original draft preparation, C.B., X.T., S.Z.; writing—review and editing, all authors; visualization, C.B., X.T.; supervision, K.P.G., A.C.D, S.B.G.; and All authors have read and agreed to the published version of the manuscript. # Authors contributed equally to the manuscript.

## Acknowledgements

We thank the DWG and the AE for providing participants, and contributions made by all participants to this research.

## Availability of data and materials

All data generated or analyzed during this study are included in this article.

## References

1. Offierski CM, and MacNab I. Hip-spine syndrome. Spine (Phila Pa 1976). 1983;8(3):316–21.

2. Chavarria JC, Douleh DG, and York PJ. The Hip-Spine Challenge. J Bone Joint Surg Am. 2021;103(19):1852–60.

3. Sing DC, Barry JJ, Aguilar TU, Theologis AA, Patterson JT, Tay BK, et al. Prior Lumbar Spinal Arthrodesis Increases Risk of Prosthetic-Related Complication in Total Hip Arthroplasty. J Arthroplasty. 2016;31(9 Suppl):227–32.e1.

4. Malkani AL, Garber AT, Ong KL, Dimar JR, Baykal D, Glassman SD, et al. Total Hip Arthroplasty in Patients With Previous Lumbar Fusion Surgery: Are There More Dislocations and Revisions? J Arthroplasty. 2018;33(4):1189–93.

5. Malkani AL, Himschoot KJ, Ong KL, Lau EC, Baykal D, Dimar JR, et al. Does Timing of Primary Total Hip Arthroplasty Prior to or After Lumbar Spine Fusion Have an Effect on Dislocation and Revision Rates? J Arthroplasty. 2019;34(5):907–11.

6. Diebo BG, Beyer GA, Grieco PW, Liu S, Day LM, Abraham R, et al. Complications in Patients Undergoing Spinal Fusion After THA. Clin Orthop Relat Res. 2018;476(2):412–7.

7. Mills ES, Bouz GJ, Formanek BG, Chung BC, Wang JC, Heckmann ND, et al. Timing of Total Hip Arthroplasty Affects Lumbar Spinal Fusion Outcomes. Clin Spine Surg. 2022;35(2):E333–e8.

8. Radcliff KE, Rihn J, Hilibrand A, DiIorio T, Tosteson T, Lurie JD, et al. Does the duration of symptoms in patients with spinal stenosis and degenerative spondylolisthesis affect outcomes?: analysis of the Spine Outcomes Research Trial. Spine (Phila Pa 1976). 2011;36(25):2197–210.

9. Tang WM, and Chiu KY. Primary total hip arthroplasty in patients with ankylosing spondylitis. J Arthroplasty. 2000;15(1):52–8.

10. Pepke W, Innmann MM, and Akbar M. [Battle: Indication for surgery in hip-spine syndrome-Hip or spine first? : The spine surgeon’s view]. Orthopade. 2020;49(10):905–12.

11. Clarius M, Farweez M, and Innmann MM. [Battle: Total hip arthroplasty or spine surgery first in patients with hip-spine-syndrome? : The arthroplasty surgeon’s point of view]. Orthopade. 2020;49(10):899–904.

12. Liu N, Goodman SB, Lachiewicz PF, and Wood KB. Hip or spine surgery first?: a survey of treatment order for patients with concurrent degenerative hip and spinal disorders. Bone Joint J. 2019;101-b(6_Supple_B):37–44.

13. Fayaz HC, Smith RM, Ebrahimzadeh MH, Pape HC, Parvizi J, Saleh KJ, et al. Improvement of Orthopedic Residency Programs and Diversity: Dilemmas and Challenges, an International Perspective. Arch Bone Jt Surg. 2019;7(4):384–96.

14. Drossard S. Structured surgical residency training in Germany: an overview of existing training programs in 10 surgical subspecialties. Innov Surg Sci. 2019;4(1):15–24.

15. Pham MH, Jakoi AM, Wali AR, and Lenke LG. Trends in spine surgery training during neurological and orthopaedic surgery residency: a 10-year analysis of ACGME case log data. JBJS. 2019;101(22):e122.

16. Burkhardt JK, Zinn PO, Bozinov O, Colen RR, Bertalanffy H, and Kasper EM. Neurosurgical education in Europe and the United States of America. Neurosurg Rev. 2010;33(4):409–17.

17. Stienen MN, Gempt J, Gautschi OP, Demetriades AK, Netuka D, Kuhlen DE, et al. Neurosurgical resident training in Germany. Journal of Neurological Surgery Part A: Central European Neurosurgery. 2017;78(04):337–43.

18. Eysenbach G. Improving the quality of Web surveys: the Checklist for Reporting Results of Internet E-Surveys (CHERRIES). J Med Internet Res. 2004;6(3):e34.

19. Rodkey DL, Lundy AE, Tracey RW, and Helgeson MD. Hip-Spine Syndrome: Which Surgery First? Clinical spine surgery. 2022;35(1):1–3.

20. Bouthors C, Benzakour A, and Court C. Surgical treatment of thoracic disc herniation: an overview. Int Orthop. 2019;43(4):807–16.

21. Deer T, Sayed D, Michels J, Josephson Y, Li S, and Calodney AK. A Review of Lumbar Spinal Stenosis with Intermittent Neurogenic Claudication: Disease and Diagnosis. Pain Med. 2019;20(Suppl 2):S32–s44.

22. Grøvle L, Haugen AJ, Keller A, Natvig B, Brox JI, and Grotle M. The bothersomeness of sciatica: patients’ self-report of paresthesia, weakness and leg pain. European Spine Journal. 2010;19:263–9.

23. Salib CG, Reina N, Perry KI, Taunton MJ, Berry DJ, and Abdel MP. Lumbar fusion involving the sacrum increases dislocation risk in primary total hip arthroplasty. Bone Joint J. 2019;101-b(2):198–206.

24. Anderson PM, Arnholdt J, and Rudert M. Total Hip Arthroplasty After Spinal Fusion Surgery. Z Orthop Unfall. 2020;158(3):333–41.

25. Oshima Y, Watanabe N, Iizawa N, Majima T, Kawata M, and Takai S. Knee-Hip-Spine Syndrome: Improvement in Preoperative Abnormal Posture following Total Knee Arthroplasty. Adv Orthop. 2019;2019:8484938.

26. Blizzard DJ, Nickel BT, Seyler TM, and Bolognesi MP. The Impact of Lumbar Spine Disease and Deformity on Total Hip Arthroplasty Outcomes. Orthop Clin North Am. 2016;47(1):19–28.

27. Clarius M, and Clarius LM. [Fast-track arthroplasty-intra- and post-operative management]. Orthopade. 2020;49(4):318–23.

