## Supplementary material for "Insights into Professional Preferences and Rationale for Surgical Sequencing in Managing Hip-Spine Syndrome": Figure S1, Table S1-6

**Supplementary materials**

**Figure S1.** The online questionnaire in English with five fictional hip-spine syndrome scenarios was previously designed by the Stanford University.
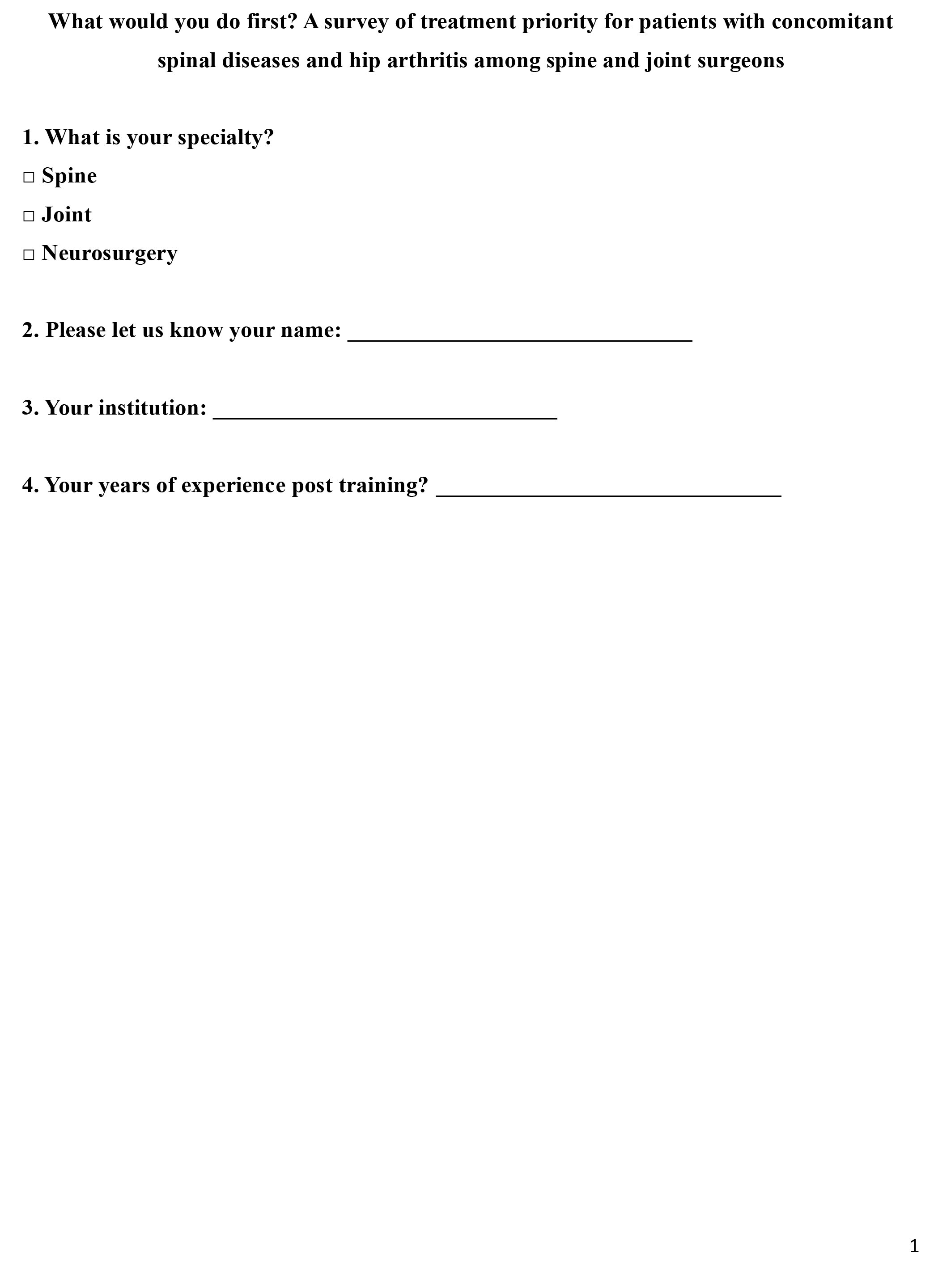

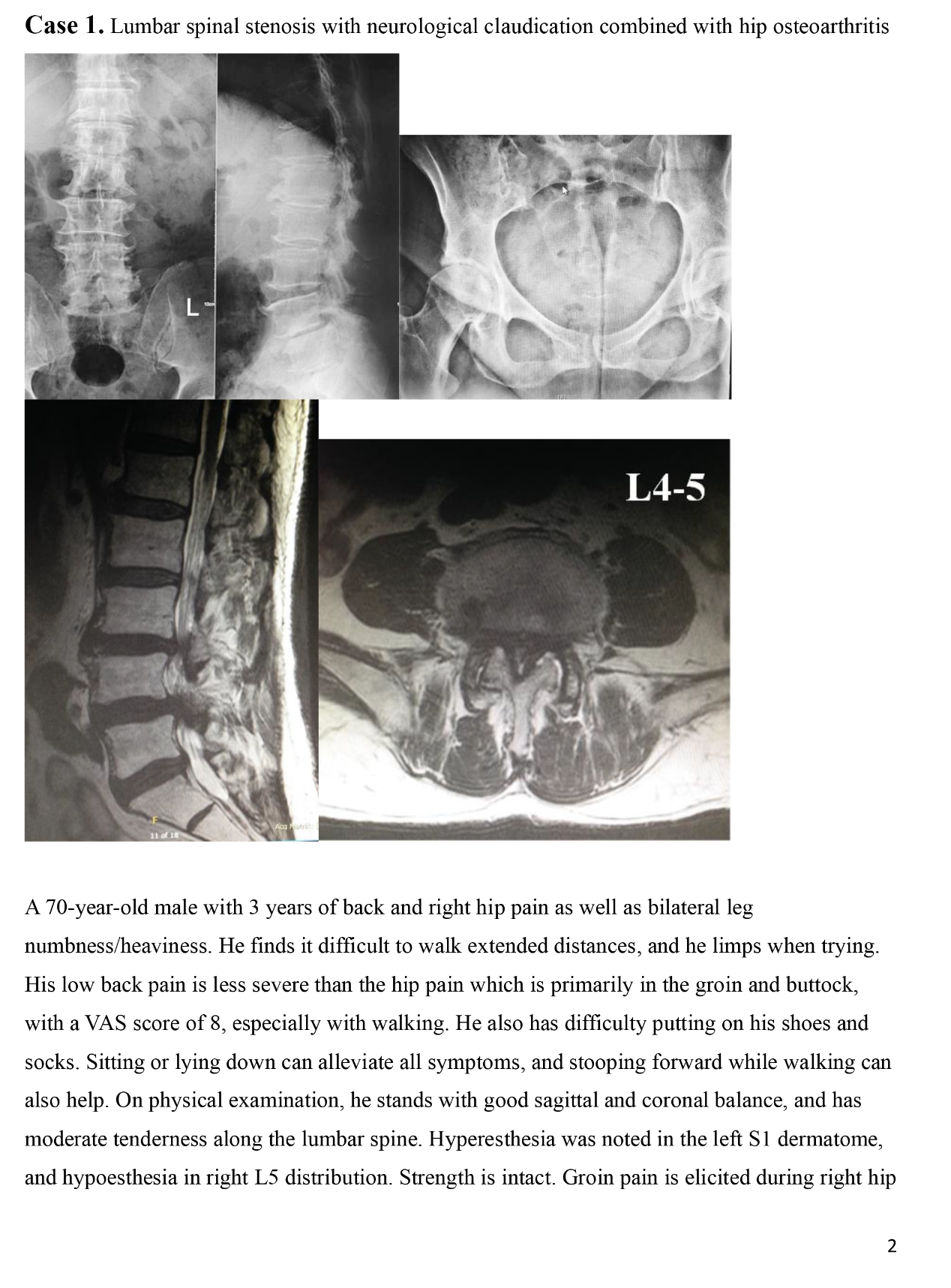

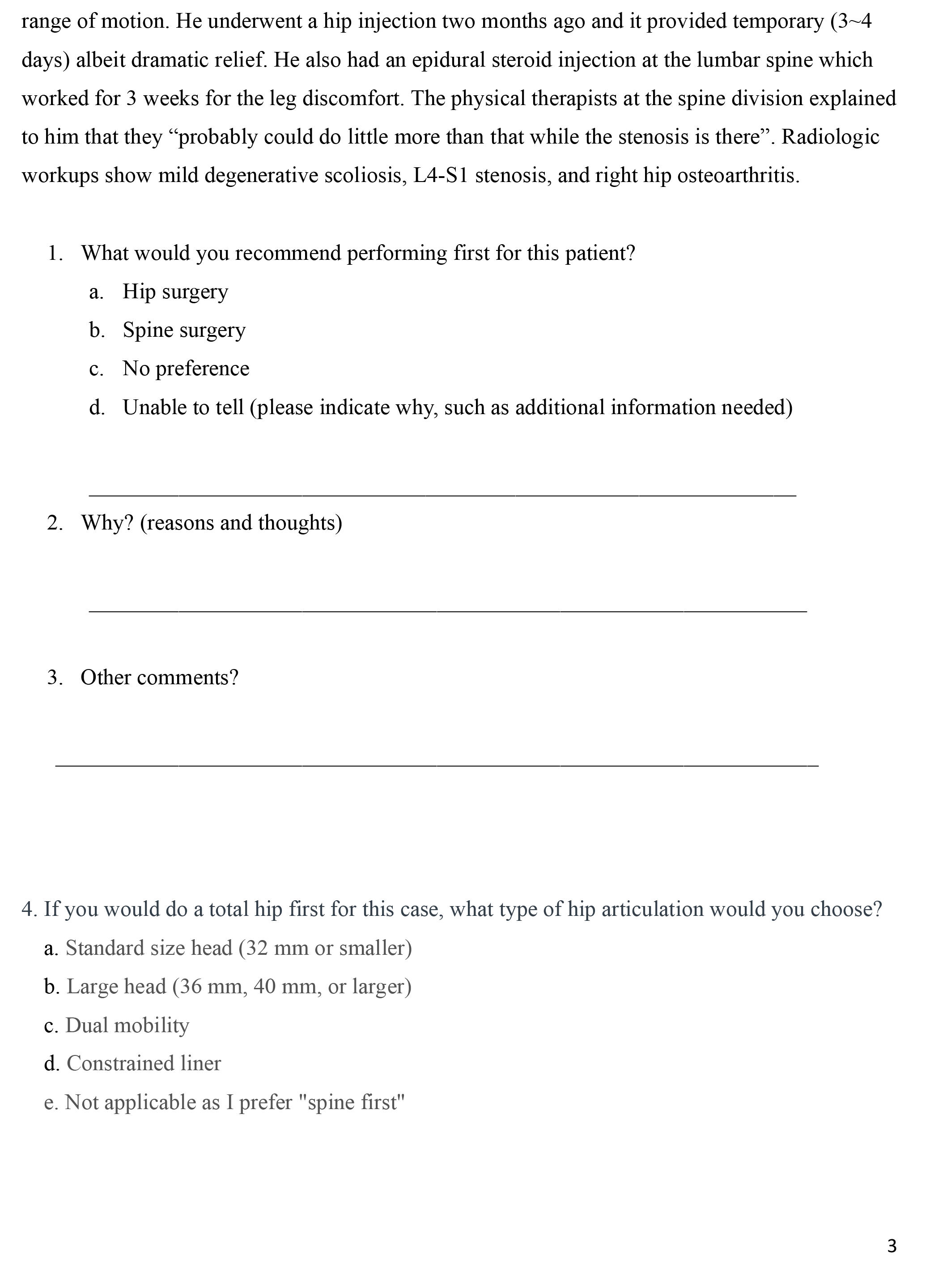

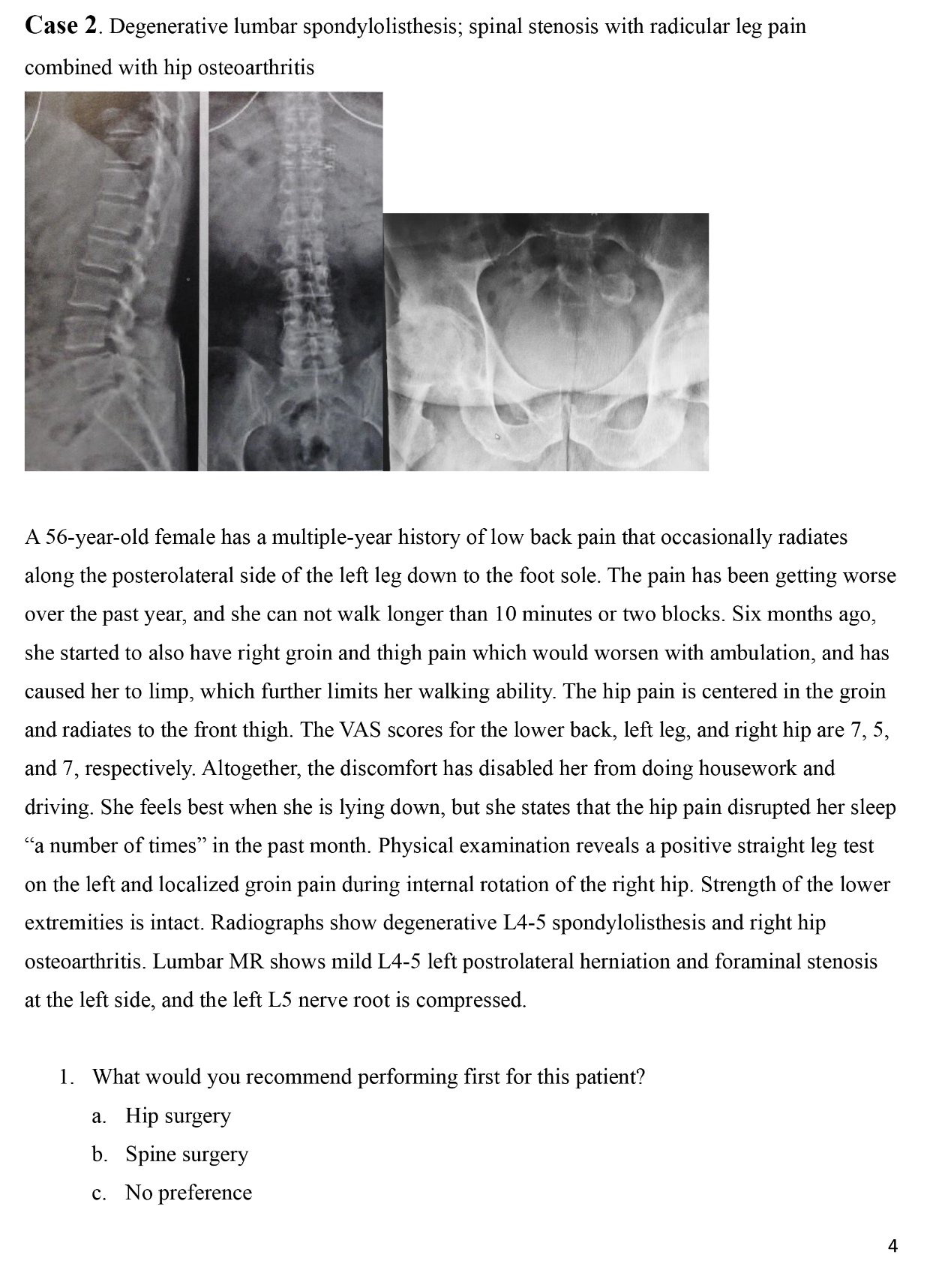

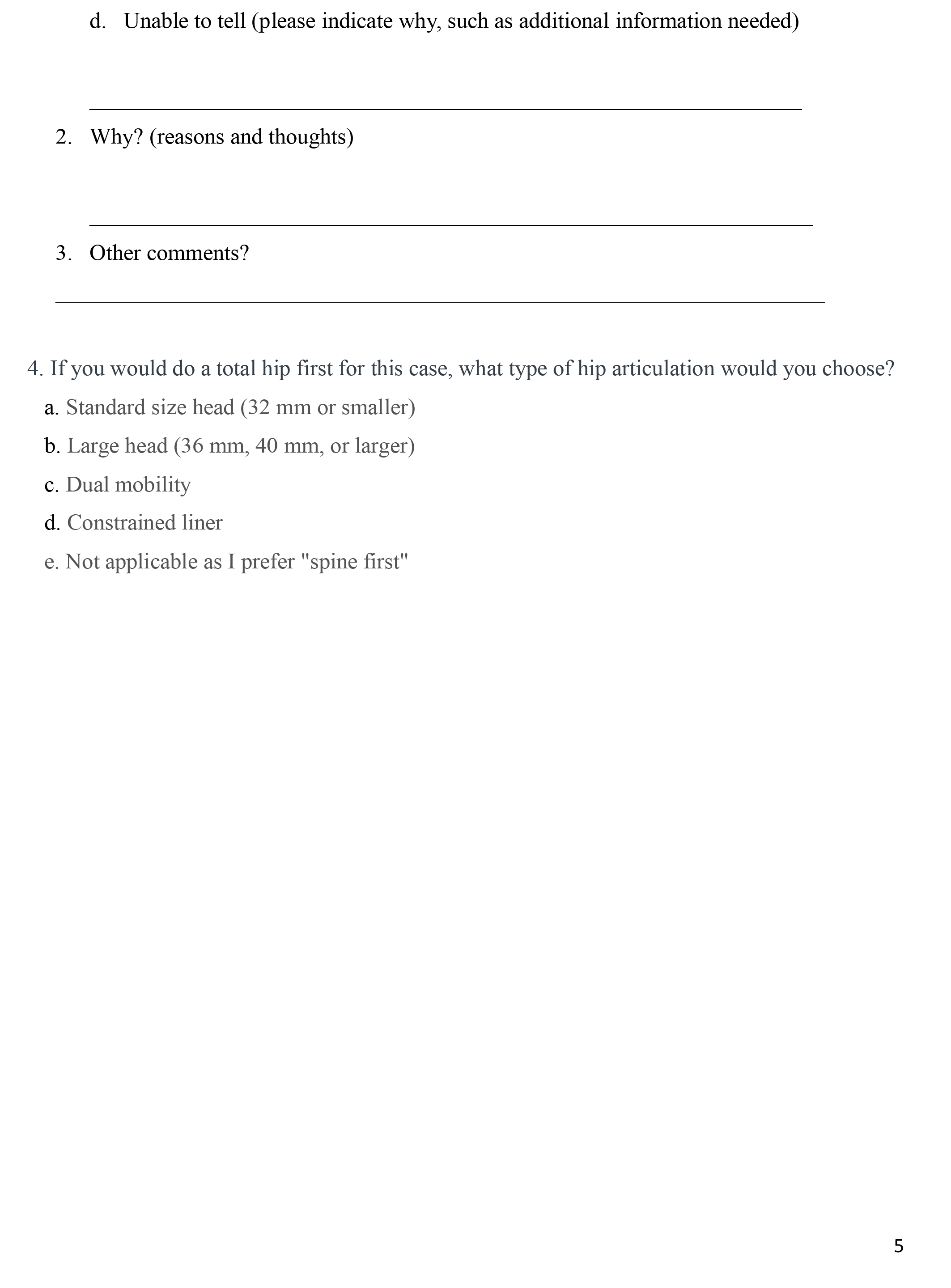

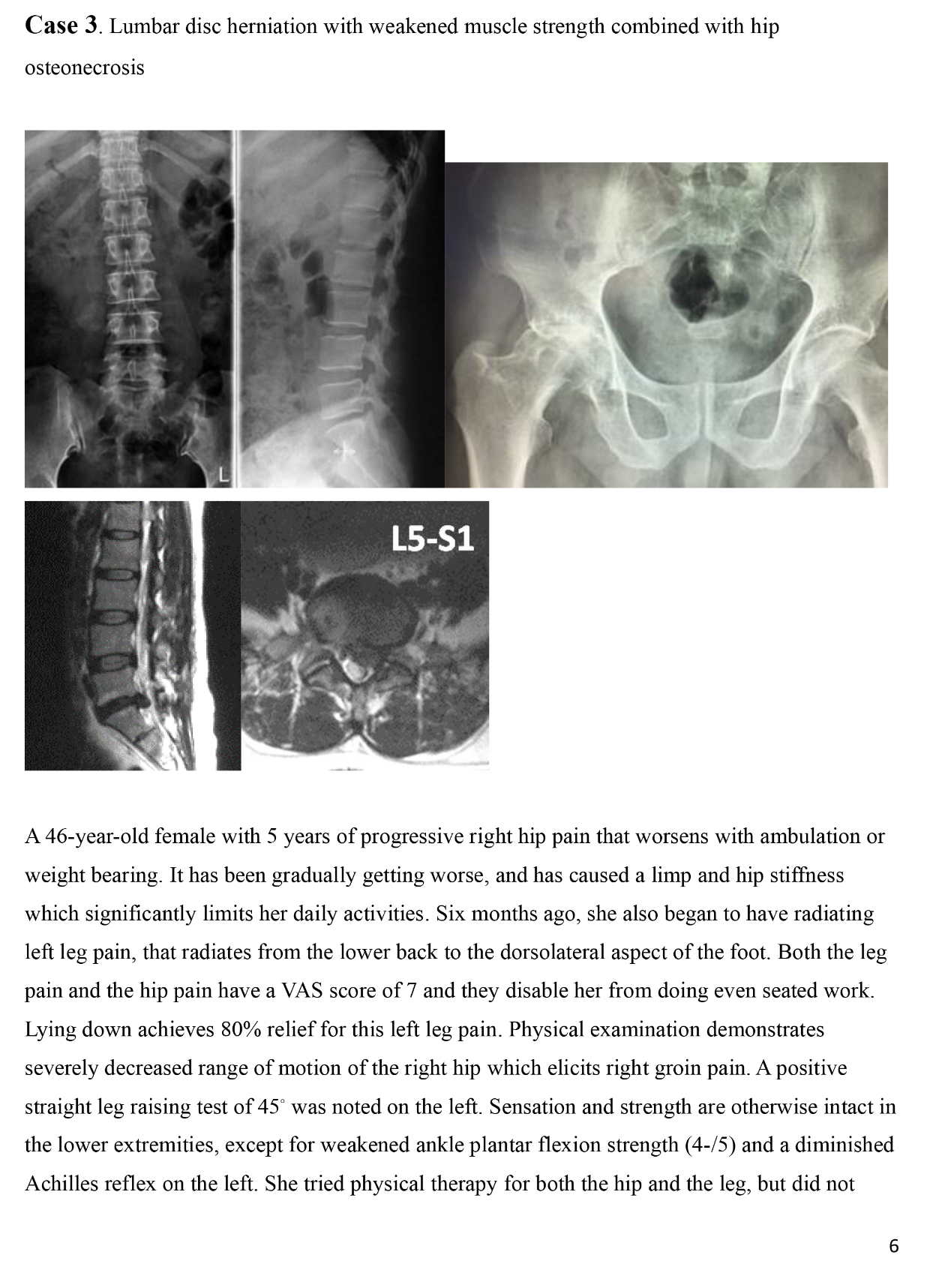

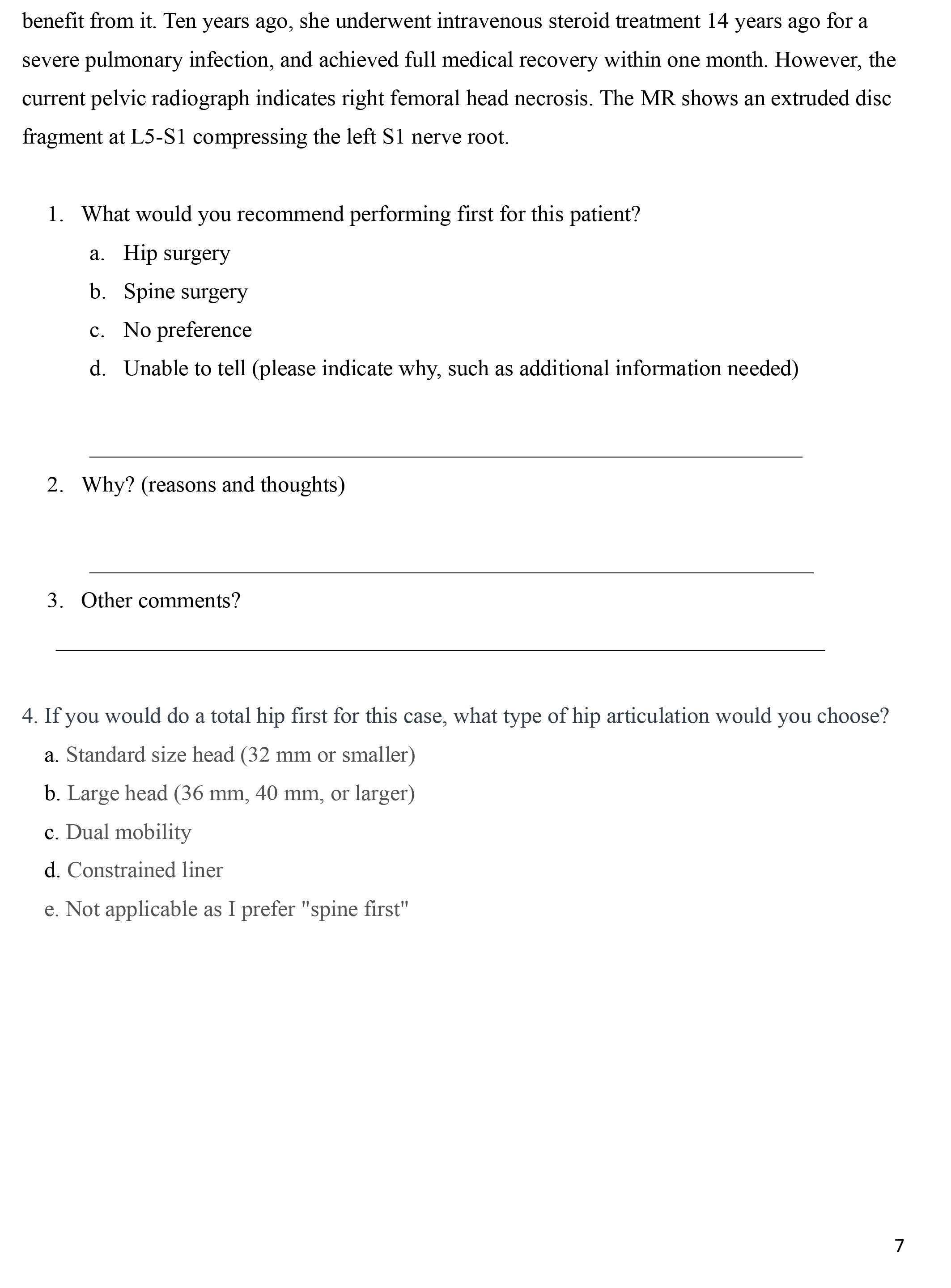

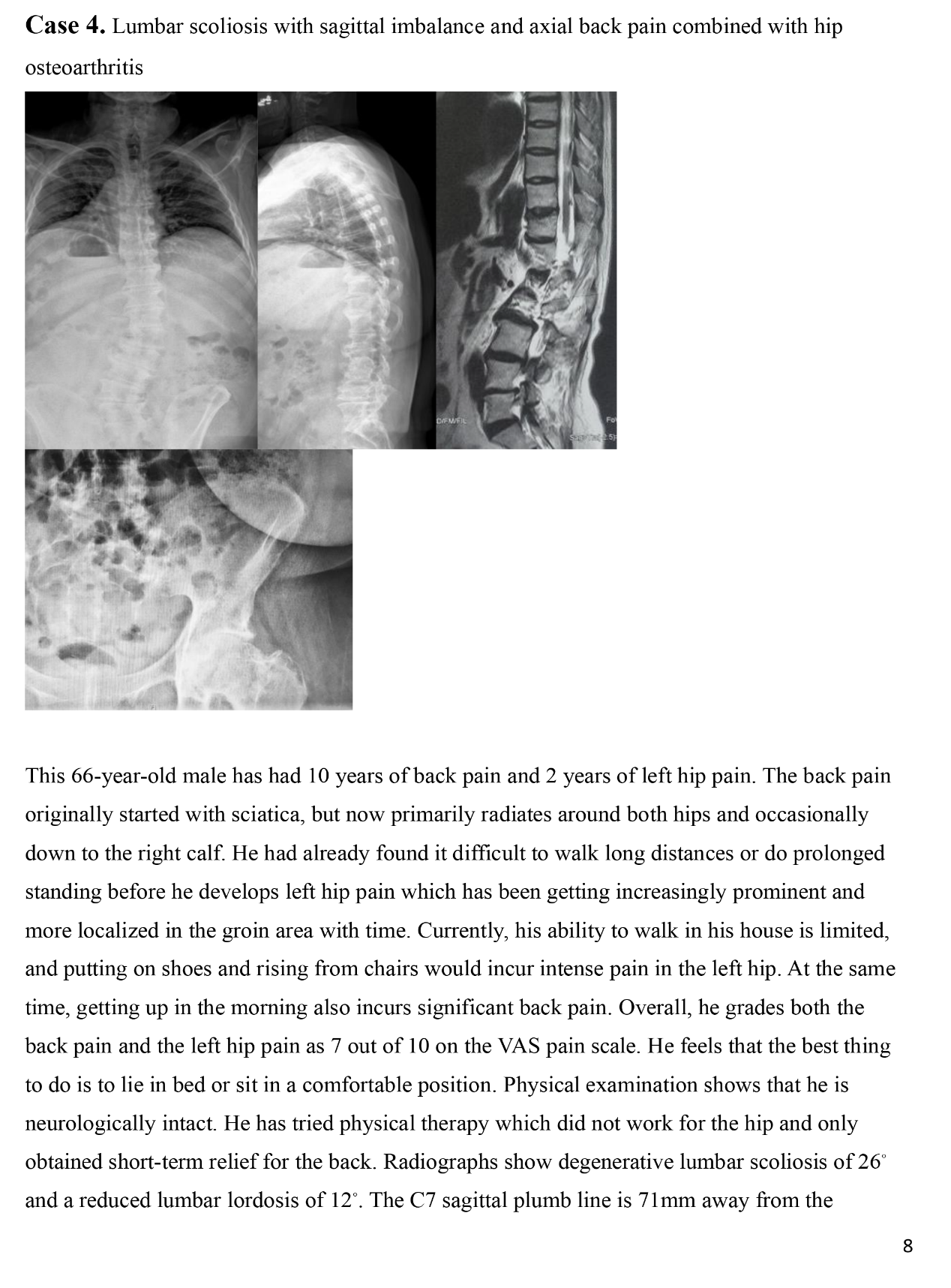

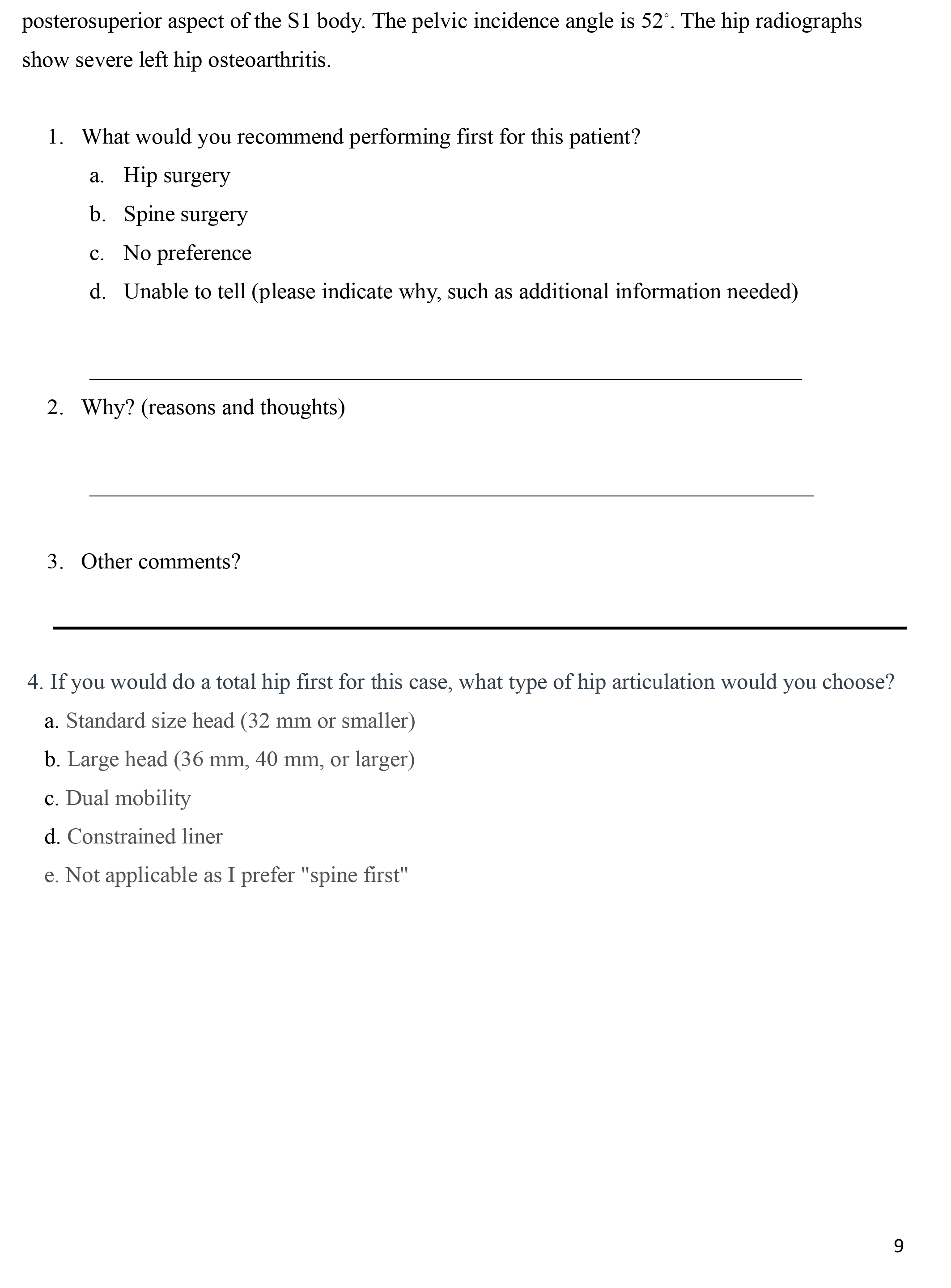

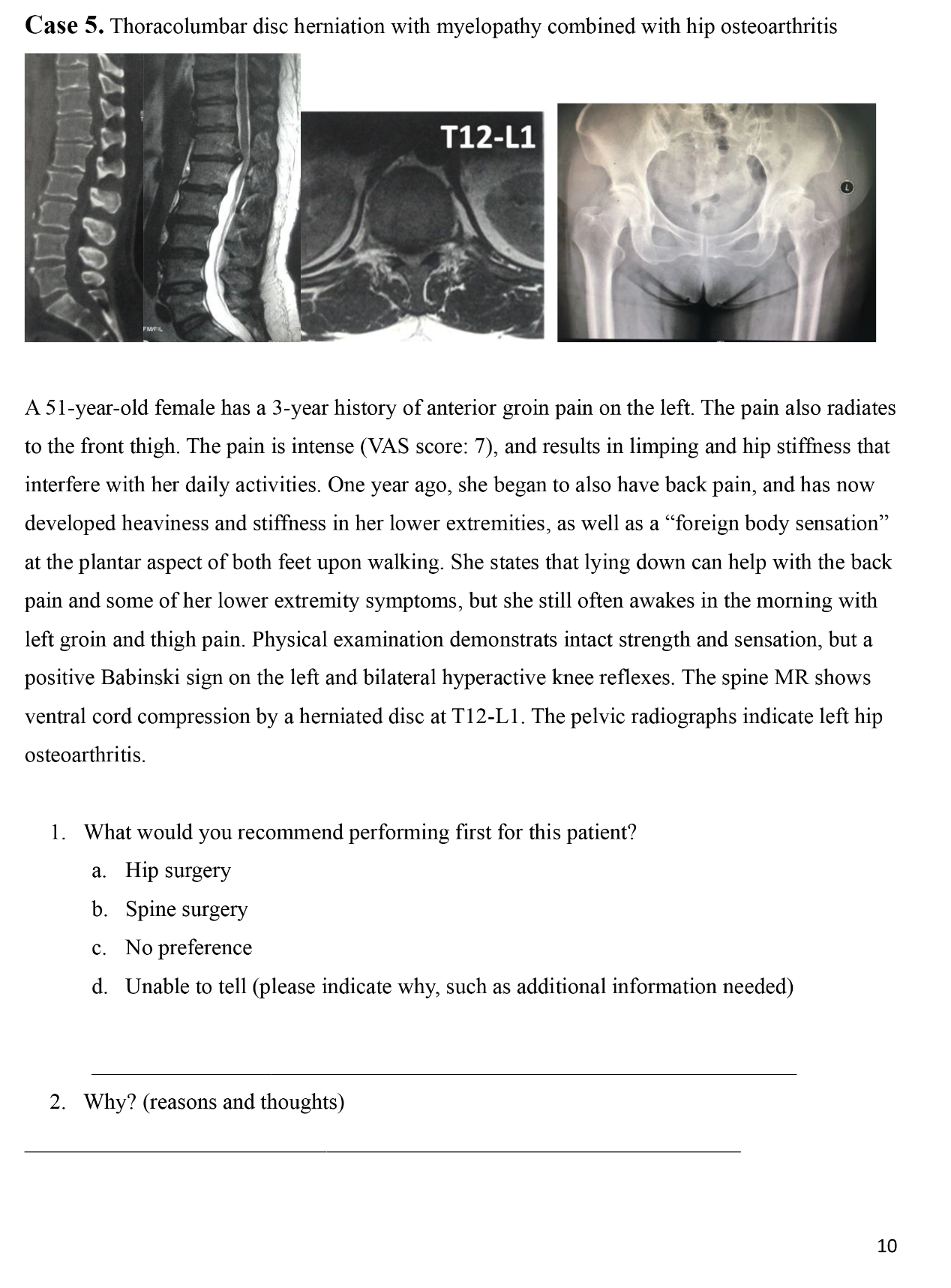

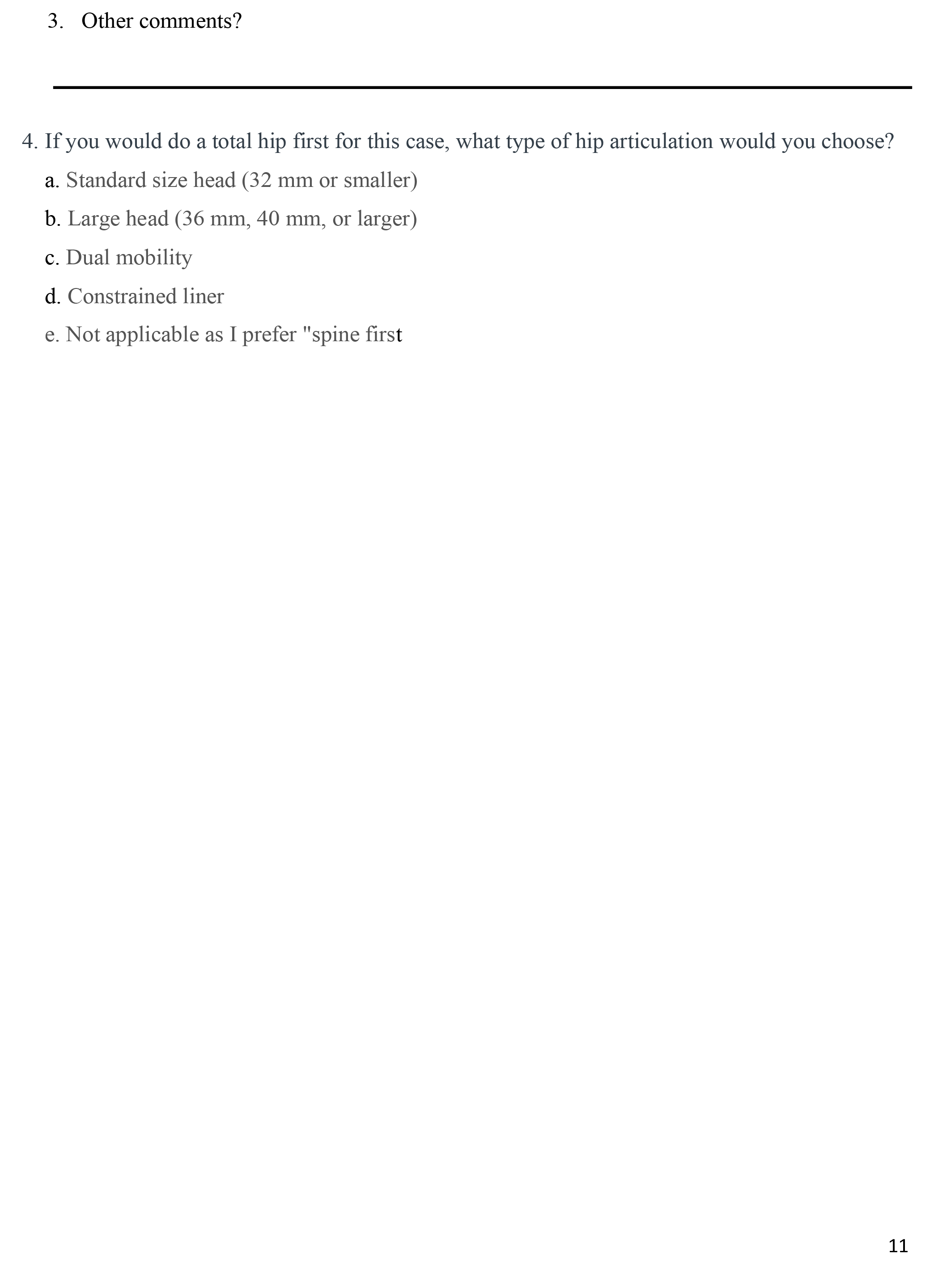

**Table S1: Checklist for Reporting Results of Internet E-Surveys (CHERRIES)**

| ***Checklist Item*** | ***Explanation*** | ***Page Number*** |
| --- | --- | --- |
| Describe survey design | The target population of the questionnaire survey is the 2,500 members of the "German Spine Association" including neurosurgical spine surgeons, and orthopedic spine surgeons, and the 883 members of the "German Joint Replacement Association" (Arbeitsgemeinschaft für Endoprothetik), mainly hip surgeons. The sample isn’t a convenience sample. | 3 |
| IRB approval | The study received ethical approval from the local ethics committee of the Technical University of Dresden. | 15 |
| Informed consent | Participants were informed by email of the approximate length time of the survey, where and when the data will be stored, who the investigator were, and the purpose of the study. | 3 |
| Data protection | Personal information in this study was only used to count questionnaire completions and to guide subsequent reminders to non-completers. | - |
| Development and testing | The electronic questionnaire for this study was pre-established by our collaborative Stanford team. Its usability and technical functionality underwent testing in one previous study, ensuring its reliability for the current investigation. | 3 |
| Open survey versus closed survey | This study was a closed survey study open only to a sample of recipients to whom the email was sent. | 3 |
| Contact mode | Initial contact with potential participants for this study was made over the Internet by sending the questionnaire via mail. | 3 |
| Advertising the survey | This survey was not conducted with any advertising or publicity. | - |
| Web/E-mail | This study sent the survey via email, with responses manually entered into a database. | 3-4 |
| Context | This study did not use any survey web sites. | - |
| Mandatory/voluntary | This was a voluntary survey. | - |
| Incentives | This survey didn’t offer any incentives. | - |
| Time/Date | The survey was conducted for 2 months. | - |
| Randomization of items or questionnaires | This study was a full-sample survey and did not require randomization. | 4 |
| Adaptive questioning | This study used adaptive questioning. | - |
| Number of Items | The number of questionnaire items per page was 4. | - |
| Number of screens (pages) | The questionnaire was distributed in 11 pages. | - |
| Completeness check | The completeness of the questionnaire was checked via "LimeSurvey" after the questionnaire had been sent. The list of participants could be filtered according to "questionnaire completed". Only the data of participants with complete questionnaires were analyzed. Each question, except the "other comments" field, had to be answered by the participants so they could only move on to the next case once they had answered the questions. | - |
| Review step | Respondents were able to review and change their answers before replying to the email. | - |
| Unique site visitor | Each participant is assigned to an IP address. | - |
| View rate (Ratio of unique survey visitors/unique site visitors) | Not applicable. | - |
| Participation rate (Ratio of unique visitors who agreed to participate/unique first survey page visitors) | Not applicable. | - |
| Completion rate (Ratio of users who finished the survey/users who agreed to participate) | The survey goas over several pages. The “completeness rate” is 30.9%. | - |
| Cookies used | No cookie was used to assign a unique user identifier to each client computer. | - |
| IP check | The client computer's IP address was not used to identify potential duplicate entries from the same user. | - |
| Log file analysis | Text-mining was used to analyze the free-text comments and to identify the most frequently used words. It was conducted with R version 4.0.4 using word frequency packages tidyverse (version 1.3.1), tidytext (version 0.3.2), wildyr, igraph and ggraph. Then, the list of the words was sorted alphabetically in Excel to summarize words with an equal meaning in main groups. For the transfer to Excel the packages readxl (version 1.3.1) and xlsx (version 0.6.5) were used. | 3 |
| Registration | This study received survey results through email responses from participants. | 3 |
| Handling of incomplete questionnaires | Only completed questionnaires were analyzed. | - |
| Questionnaires submitted with an atypical timestamp | The survey time per participant was not measured. No time frame has been set. | - |
| Statistical correction | None of these methods were used. | - |

This checklist has been modified from Eysenbach G. Improving the quality of Web surveys: the Checklist for Reporting Results of Internet E-Surveys (CHERRIES). J Med Internet Res. 2004 Sep 29;6(3):e34 [erratum in J Med Internet Res. 2012; 14(1): e8.]. Article available at [https://www.jmir.org/2004/3/e34](https://www.jmir.org/2004/3/e34/)/; erratum available <https://www.jmir.org/2012/1/e8/>. Copyright ©Gunther Eysenbach. Originally published in the [Journal of Medical Internet](http://www.jmir.org) Research, 29.9.2004 and 04.01.2012.

This is an open-access article distributed under the terms of the Creative Commons Attribution License (<https://creativecommons.org/licenses/by/2.0/>), which permits unrestricted use, distribution, and reproduction in any medium, provided the original work, first published in the Journal of Medical Internet Research, is properly cited.

**Table. S2. Sequence selection and comparison of surgical procedures by different specialists in Germany at different year of experience levels in scenario 1.**

| **Case 1^a^** | | | | | |
| --- | --- | --- | --- | --- | --- |
| Years of experience | Specialty | Hip-first (%) | Spine-first (%) | No preference (%) | χ-value; p-value |
| 0-10 years | Hip surgeon | 53.8 | 46.2 | 0 | 6.485; 0,166 |
|  | Neurosurgical spine surgeon | 0 | 80 | 20 |  |
|  | Orthopaedic spine surgeon | 47.8 | 47.8 | 4.4 |  |
| 11-20 years | Hip surgeon | 31.8 | 45.5 | 22.7 | 11.943; 0,018* |
|  | Neurosurgical spine surgeon | 11.8 | 88.2 | 0 |  |
|  | Orthopaedic spine surgeon | 40.9 | 54.5 | 4.6 |  |
| 21-30 years | Hip surgeon | 38.9 | 61.1 | 0 | 7.104; 0.130 |
|  | Neurosurgical spine surgeon | 7.7 | 84.6 | 7.7 |  |
|  | Orthopaedic spine surgeon | 50 | 41.7 | 8.3 |  |
| > 30 years | Hip surgeon | 11.1 | 66.7 | 22.2 | 8.089; 0.088 |
|  | Neurosurgical spine surgeon | 0 | 100 | 0 |  |
|  | Orthopaedic spine surgeon | 100 | 0 | 0 |  |

^a^ Lumbar canal stenosis with neurogenic claudication combined with osteoarthritis of the hip

**Table. S3. Sequence selection and comparison of surgical procedures by different specialists in Germany at different year of experience levels in scenario 2.**

| **Case 2^b^** | | | | | |
| --- | --- | --- | --- | --- | --- |
| Years of experience | Specialty | Hip-first (%) | Spine-first (%) | No preference (%) | χ-value; p-value |
| 0-10 years | Hip surgeon | 84.6 | 15.4 | 0 | 5.586; 0.232 |
|  | Neurosurgical spine surgeon | 80 | 20 | 0 |  |
|  | Orthopaedic spine surgeon | 91.3 | 0 | 8.7 |  |
| 11-20 years | Hip surgeon | 91 | 4.5 | 4.5 | 2.243; 0.691 |
|  | Neurosurgical spine surgeon | 76.4 | 11.8 | 11.8 |  |
|  | Orthopaedic spine surgeon | 91 | 4.5 | 4.5 |  |
| 21-30 years | Hip surgeon | 83.3 | 11.1 | 5.6 | 15.663; 0.004* |
|  | Neurosurgical spine surgeon | 38.5 | 7.7 | 53.8 |  |
|  | Orthopaedic spine surgeon | 91.7 | 8.3 | 0 |  |
| > 30 years | Hip surgeon | 100 | 0 | 0 | 5.833; 0,212 |
|  | Neurosurgical spine surgeon | 50 | 25 | 25 |  |
|  | Orthopaedic spine surgeon | 100 | 0 | 0 |  |

^b^ Degenerative lumbar spondylolisthesis with radicular leg pain combined with osteoarthritis of the hip

**Table. S4. Sequence selection and comparison of surgical procedures by different specialists in Germany at different year of experience levels in scenario 3.**

| **Case 3^c^** | | | | | |
| --- | --- | --- | --- | --- | --- |
| Years of experience | Specialty | Hip-first (%) | Spine-first (%) | No preference (%) | χ-value; p-value |
| 0-10 years | Hip surgeon | 38.5 | 53.8 | 7.7 | 3.151; 0.533 |
|  | Neurosurgical spine surgeon | 0 | 80 | 20 |  |
|  | Orthopaedic spine surgeon | 39.1 | 52.2 | 8.7 |  |
| 11-20 years | Hip surgeon | 45.5 | 40.9 | 13.6 | 1.882; 0.757 |
|  | Neurosurgical spine surgeon | 41.2 | 52.9 | 5.9 |  |
|  | Orthopaedic spine surgeon | 31.8 | 59.1 | 9.1 |  |
| 21-30 years | Hip surgeon | 38.9 | 55.6 | 5.5 | 4.507; 0.342 |
|  | Neurosurgical spine surgeon | 30.8 | 53.8 | 15.4 |  |
|  | Orthopaedic spine surgeon | 16.7 | 83.3 | 0 |  |
| > 30 years | Hip surgeon | 44.5 | 44.5 | 11 | 1.593; 0.810 |
|  | Neurosurgical spine surgeon | 50 | 50 | 0 |  |
|  | Orthopaedic spine surgeon | 0 | 100 | 0 |  |

^c^ Lumbar disc herniation with muscle strength weakness combined with osteoarthritis of the hip

**Table. S5. Sequence selection and comparison of surgical procedures by different specialists in Germany at different year of experience levels in scenario 4.**

| **Case 4^d^** | | | | | |
| --- | --- | --- | --- | --- | --- |
| Years of experience | Specialty | Hip-first (%) | Spine-first (%) | No preference (%) | χ-value; p-value |
| 0-10 years | Hip surgeon | 61.5 | 15.4 | 23.1 | 1.567; 0.815 |
|  | Neurosurgical spine surgeon | 80 | 0 | 20 |  |
|  | Orthopaedic spine surgeon | 69.6 | 17.4 | 13 |  |
| 11-20 years | Hip surgeon | 72.7 | 9.1 | 18.2 | 5.748; 0;219 |
|  | Neurosurgical spine surgeon | 76.5 | 23.5 | 0 |  |
|  | Orthopaedic spine surgeon | 77.3 | 18.2 | 4.5 |  |
| 21-30 years | Hip surgeon | 77.8 | 22.2 | 0 | 7.577; 0.108 |
|  | Neurosurgical spine surgeon | 61.5 | 7.7 | 30.8 |  |
|  | Orthopaedic spine surgeon | 75 | 16.7 | 8.3 |  |
| > 30 years | Hip surgeon | 66.7 | 33.3 | 0 | 0,525; 0,769 |
|  | Neurosurgical spine surgeon | 75 | 25 | 0 |  |
|  | Orthopaedic spine surgeon | 100 | 0 | 0 |  |

^d^ Scoliosis with back pain combined with osteoarthritis of the hip

**Table. S6. Sequence selection and comparison of surgical procedures by different specialists in Germany at different year of experience levels in scenario 5.**

| **Case 5^e^** | | | | | |
| --- | --- | --- | --- | --- | --- |
| Years of experience | Specialty | Hip-first (%) | Spine-first (%) | No preference (%) | χ-value; p-value |
| 0-10 years | Hip surgeon | 0 | 84.6 | 15.4 | 1.941; 0,379 |
|  | Neurosurgical spine surgeon | 0 | 100 | 0 |  |
|  | Orthopaedic spine surgeon | 0 | 95.7 | 4.3 |  |
| 11-20 years | Hip surgeon | 10.5 | 86.4 | 4.5 | 5,593; 0,232 |
|  | Neurosurgical spine surgeon | 0 | 100 | 0 |  |
|  | Orthopaedic spine surgeon | 0 | 100 | 0 |  |
| 21-30 years | Hip surgeon | 5.6 | 94.4 | 0 | 3.762; 0.439 |
|  | Neurosurgical spine surgeon | 0 | 100 | 0 |  |
|  | Orthopaedic spine surgeon | 8.3 | 83.4 | 8.3 |  |
| > 30 years | Hip surgeon | 11.1 | 88.9 | 0 | 3.144; 0.534 |
|  | Neurosurgical spine surgeon | 0 | 75 | 25 |  |
|  | Orthopaedic spine surgeon | 0 | 100 | 0 |  |

^e^ Thoracolumbar disc herniation with myelopathy combined with osteoarthritis of the hip
